# Exploring Cardiometabolic Health, Physical Activity, and Quality of Life in Patients with Obesity and Multimorbidity during an Intensive Weight Loss Program: A Protocol for a Real-Life Prospective Cohort Study

**DOI:** 10.1101/2025.10.20.25337060

**Authors:** Rasmus Hoxer Brødsgaard, Carsten Dirksen, Mette Merete Pedersen, Jeanette Wassar Kirk, Olivia Maltha Duun, Minna Titika Finken, Kirstine Nyvold Bojsen-Møller, Thomas Bandholm

## Abstract

**Background:** Obesity is associated with increased risk of morbidity, reduced quality of life, and pervasive weight stigma. For patients with severe obesity and multimorbidity who are ineligible for bariatric surgery, effective treatment options remain limited. Recently, glucagon-like peptide 1 receptor agonist-based weight loss medications have shown promise in weight management, but their effects in highly complex patient populations are not well described.

**Objective:** This protocol outlines a prospective observational study designed to explore and describe changes in cardiometabolic health, physical activity, physical capacity, and wellbeing among patients with obesity and multimorbidity enrolled in a two-year intensive weight loss program with GLP-1RA medication.

**Methods:** The study uses an exploratory prospective longitudinal cohort design and is based on an intensive weight loss program for patients with obesity and multimorbidity at Copenhagen University Hospital - Hvidovre. Approximately 35 patients will be enrolled over a six-month period. Data are collected at baseline, four months, and 24 months. Key outcomes include objectively measured physical activity, health-related quality of life, and 10-year cardiovascular risk. Additional outcomes include anthropometrics, mental health-related quality of life, weight bias internalization, cardiorespiratory fitness, lower body strength, and walking capacity. Data will be analyzed descriptively, and changes will be reported as mean differences with 95% confidence intervals. The study was registered at clinicaltrials.gov prior to recruitment (NCT06234111).

**Perspective:** This study will provide novel insights into the health effects of intensive GLP-1RA-based weight loss program for patients with severe obesity and multimorbidity ineligible for bariatric surgery, thereby addressing an important knowledge gap in obesity management.

**Trial Registration:** The study has been pre-registered with clinicaltrials.gov prior to inclusion of the first participant (<u>NCT06234111</u>).

**Project group:** *SemPAct project group:* Rasmus H. Brødsgaard (principal investigator), Thomas Bandholm, Kirstine N. Bojsen-Møller, Carsten Dirksen, Jeanette W. Kirk, Mette M. Pedersen, Sara O. Hofmann, Emilie S. Larsen, Minna Titika Finken, and Olivia M. Duun.

## Background and Rationale

More and more people struggle with the numerous and serious consequences of obesity (1,2). It poses significant health risks (3,4) and fosters pervasive weight stigma that negatively impacts mental health and well-being (5–7). Moreover, obesity is more common in individuals with low physical activity levels (8) and lower socioeconomic status (9,10). While weight loss can improve cardiometabolic health in individuals with obesity (11,12), traditional weight loss approaches often yield only short-term results (13), which may be explained in part by a tendency towards reduced physical activity after weight loss (14,15). Bariatric surgery offers significant and long-term weight loss for, but only for individuals with severe degrees of obesity and established obesity-related comorbidity (16). Furthermore, there are a subpopulation of patients with obesity who fulfil the eligibility criteria for bariatric surgery but are ineligible for surgery due to living with severe somatic or psychiatric illness (17).

Recently, the new class of weight loss medication, the glucagon-like-peptide 1 receptor agonists (GLP-1RAs), appears to be revolutionizing obesity management. GLP-1RAs, such as Semaglutide, have proven highly effective in long-term weight loss maintenance (18), but only for as long as the treatment is continued (19). Therefore, GLP-1RAs are not recommended as a stand-alone solution but should be accompanied by lifestyle changes such as healthy diets and physical activity (12). Specifically, physical activity and exercise has been found to play a pivotal role in enhancing the health of individuals using GLP-1RAs (20) and at preventing the otherwise expected weight regain after discontinuation of the medication (21). Consequently, GLP-1RA-based lifestyle interventions are a welcome strategy for weight management, especially for patients with more severe levels of obesity and multiple chronic conditions who are expected to gain tangible and significant health benefits from a weight loss, but for whom losing weight is particularly challenging (22,23).

At the recently established unit for “Obesity and Nutrition” based at Copenhagen University Hospital - Hvidovre, we help patients with obesity and multimorbidity who, for various reasons, are ineligible for bariatric surgery. We offer these patients a two-year highly specialized interdisciplinary intensive weight loss program consisting of GLP-1RA weight loss medication (Semaglutide) and intensive dietetic and behavioral support (24). Based on preliminary data, the program is not only effective at weight reduction; it appears to have enabled several patients to achieve substantial enhancements in their other morbidities (e.g., enhanced cardiac and pulmonary function) and has even made some patients eligible for organ transplants. However, a systematic evaluation of the program’s impact on other health outcomes, such as cardiometabolic risk factors, physical activity, physical capacity, and well-being, has yet to be conducted.

Large-scale pharmaceutical trials have demonstrated the effectiveness of GLP-1RAs in reducing weight and disease-specific symptoms and risks in populations with obesity and comorbidities, such as type 2 diabetes (25), cardiovascular disease (26), and knee osteoarthritis (27). Similarly, a few observational studies have found results of GLP-1RA weight loss programs in real-world clinical settings that are of comparable sizes to those in the major RCTs (28,29). However, none of these studies focused on patients with complex obesity and multiple debilitating chronic conditions that are ineligible for bariatric surgery which highlights the need to investigate this particularly vulnerable and vastly understudied group of patients.

The aim of this prospective observational study is, therefore, to explore and describe changes in cardiometabolic health, physical activity, physical capacity, and wellbeing during an interdisciplinary intensive weight loss program with GLP-1RA medication in patients with obesity and multimorbidity.

## Study design

This study is part of the project “*Semaglutide and Physical Activity for Obesity and Multimorbidity: Co-designing Healthy Healthcare”*. The overall goal of the project is to support patients with obesity and multimorbidity in an intensive weight loss program to adopt healthier, more physically active lifestyles, thereby reducing risks of obesity-related diseases and improving general health. To achieve this goal, we will conduct three studies:

1. A *prospective cohort study* to explore and describe changes in patients’ cardiometabolic health, physical activity, physical capacity, and wellbeing during the weight loss program.
2. A *qualitative interview study* to explore patients’ perspectives on obesity and multimorbidity-related challenges towards physical activity (30).
3. A *co-design study* to describe the process of co-designing a tool to support patients with obesity and multimorbidity using GLP-1RA weight loss medication in engaging in physical activity.

This study protocol is, however, exclusively for the prospective cohort study. It uses an exploratory prospective longitudinal cohort design and is based on patients with obesity and multimorbidity enrolled in the intensive weight loss program at Copenhagen University Hospital - Hvidovre. The study is exploratory and descriptive and, therefore, does not include hypothesis testing. It was registered with clinicaltrials.gov prior to including the first patient (<u>NCT06234111</u>). Reporting of the study findings will adhere to the STROBE checklist using the extension for cohort studies (https://www.strobe-statement.org/).

### Setting – Obesity and Nutrition

The study is based in the unit for Obesity and Nutrition at Copenhagen University Hospital - Hvidovre. Obesity and Nutrition provides a two-year highly specialized interdisciplinary intensive weight loss program for patients with complex obesity and multimorbidity. The program’s purpose is to support patients in achieving a clinically relevant weight loss (in practice > 10%) and, thereby, reduce their morbidity burden and improve their general physical and mental health. For a detailed description of the contents of the weight loss program, please see the section “Exposure”.

The program is offered to individuals living in the Capital Region of Denmark who fulfill the criteria for bariatric surgery (17) but are considered ineligible for surgery, typically due to severe comorbidities that elevate the risk of peri- or postoperative complications. Consequently, most patients in the weight loss program are severely burdened by high levels of obesity (usually BMI > 40 kg/m^2^), have debilitating multimorbidity (such as severe organ failure or psychiatric disease), and have functional limitations in their daily activities (corresponding to Edmonton Obesity Staging System stages 2-4 (31)). In this group of patients, substantial weight loss is expected to lead to tangible and significant health benefits, even within a relatively short period (< two years). However, this has not yet been confirmed in clinical trials.

The program includes approximately 75 new patients each year. After four months in the program, the patients’ early results, including weight, are evaluated. Patients with clinically relevant weight loss (≥ 10 % reduction in body weight relative to their baseline weight) are offered to continue in the program. Currently, more than 90 % of patients meet this criterion. The few patients who do not respond to the program after four months are not offered to continue. Instead, they are referred to their general practitioner or other healthcare professionals for continuous management of their comorbidities.

By the end of the two-year program, we prepare individualized plans for continued weight loss maintenance, including plans for healthy dietary habits and for continuing or discontinuing GLP-1RA. However, it is likely that many patients will stop using GLP-1RAs at some point after having completed the program, increasing their risk of weight regain and, thereby, diminish their health benefits. Therefore, we are currently developing an exit strategy that we will design in collaboration with patients and, if sufficient funding is acquired, test it for feasibility and efficiency. The exit strategy will consist of tapering of GLP-1RA and individualized guidance on physical activity and healthy dietary habits to prevent the potential negative consequences of GLP-1RA discontinuation.

### Eligibility

Study participants are recruited from the intensive weight loss program in Obesity and Nutrition. Hence, the study uses the same set of inclusion criteria:

- Fulfilment of criteria for bariatric surgery (BMI ≥ 35 kg/m^2^, aged ≥ 18 years, and one or more obesity-related conditions (17)).
- Presence of comorbidities that prevent bariatric surgery.

Exclusion criteria are:

- Unwillingness to participate in the study.
- Inability to understand Danish.
- Cognitive impairments.

### Exposure – The Weight Loss Program

Participation in the intensive weight loss program is initiated after consultations with a physician (specialist in endocrinology) and a dietician to establish the patient’s eligibility. The program spans over two years for each patient and consists of total diet replacement, weight loss medication, and dietetic behavioral support. Diet replacement products and weight loss medication are provided free of charge throughout the entire program. The basic structure of the weight loss program is outlined in Figure 1 (leftmost column, Weight loss program activities).

**FIGURE 1:**
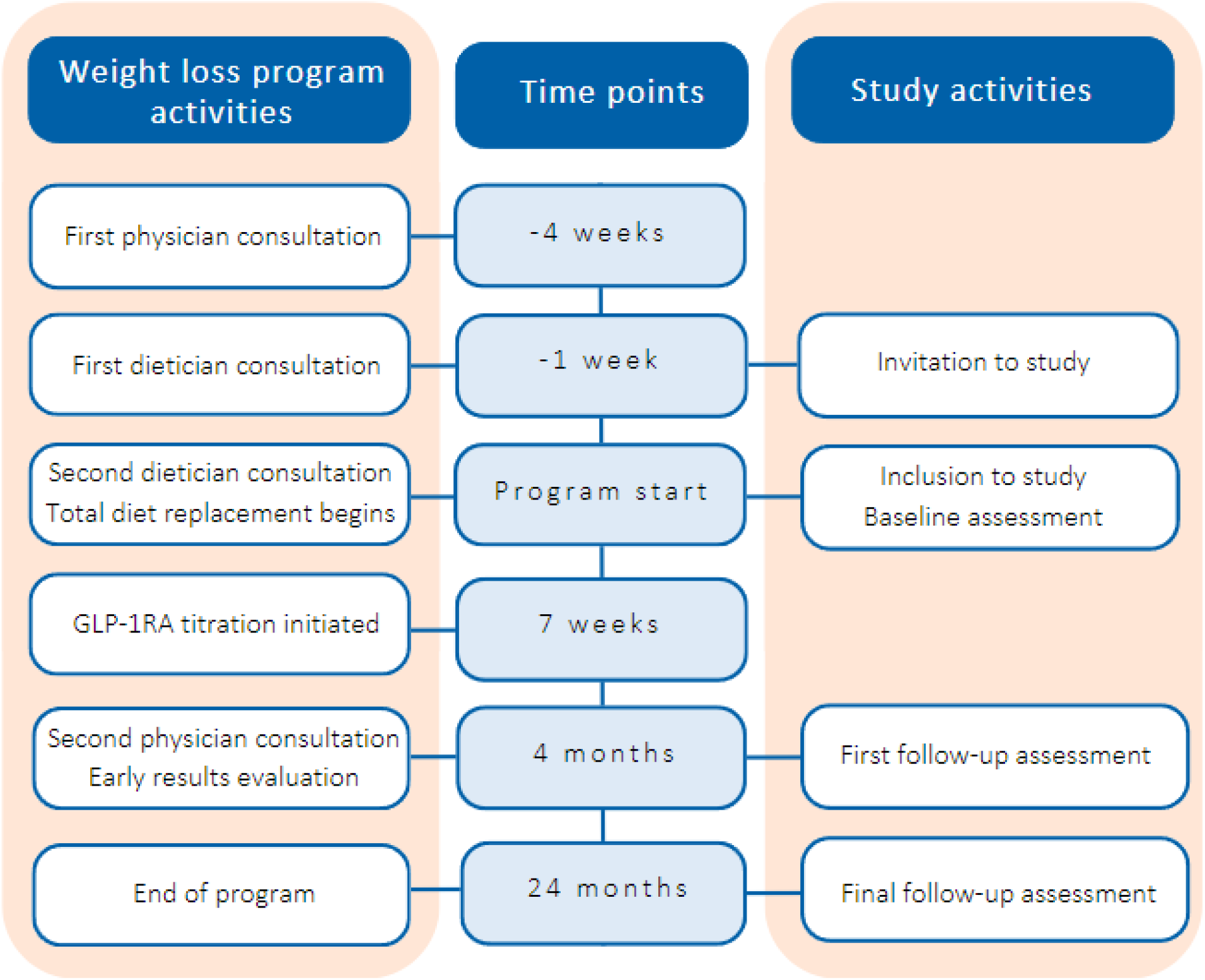
Chronological overview of the weight loss program and study activities. The leftmost column outlines the structure of program activities for all patients in the weight loss program, whereas the rightmost column outlines additional study activities only for patients included in this study.

#### Total diet replacement (TDR)

In the program’s first eight weeks, patients will replace usual diet with hypocaloric (800 kcal/day) TDR products (Nupo ApS, Brøndby, Denmark). Patients are allowed to supplement TDR with leafy vegetables to improve tolerability. After week eight, TDR is gradually reduced and replaced with a healthy calorie-reduced diet plan. The program is designed to have a highly individualized approach, allowing alterations to the standard use of TDR (e.g., additional periods of TDR).

#### Weight loss medication

After approximately seven weeks, once-weekly injections Semaglutide (Wegovy®) is initiated. The patients will follow a titration plan prescribed by the physician and attend regular consultations with a nurse, who instructs on the correct use and titration of the medication. The titration plan aims for a dose of 2.4 mg/week but accepts lower doses if adverse effects are intolerable. Once fully titrated (usually occurs after five to six months), patients will continue using the medication until the final physician consultation at month 21 of the program, where a plan is made for gradually discontinuing the weight loss medication.

#### Dietetic behavioral support

Patients attend individual dietician consultations throughout the entire program with varying frequency (biweekly in the beginning, bimonthly towards the end). In these, dieticians provide behavioral strategies to identify and manage existing maladaptive eating patterns and establish sustainable healthier eating habits. Additionally, dieticians will advise patients to gradually increase physical activity to support and enhance the health benefits of the weight loss.

### Outcomes

This observational exploratory study will explore and describe potential changes in health outcomes during the intensive weight loss program (from baseline to four months and 24 months) and not test predefined hypotheses about causal relationships between exposures and outcomes. Therefore, it is designed with a flat non-hierarchical outcome structure and multiple evenly valued outcomes measures. However, a few outcomes were identified as being of particular interest for patients and clinicians and are labelled as “key outcomes” (for further details, please see the “Patient and Public Involvement” section). Outcomes and outcome measures are detailed below.

#### Key outcomes

##### Physical activity

We will use SENS Motion® accelerometry to measure physical activity behavior for seven full days (SENS Innovation ApS, Copenhagen, Denmark). The SENS Motion® accelerometer captures detailed measurements of a range of different physical activity intensities. For the sake of simplicity, we will dichotomize its measurements into time spent physically active (standing, walking, running, and cycling) and inactive (sitting, lying down, and sleeping), focusing on the mean time/day spent physically active.

##### Health-related quality of life – physical

We will administer the 36-Item Short Form Survey questionnaire for all patients. This widely used tool has been validated in populations with obesity (32). Furthermore, it has been translated and validated into Danish (33,34). Possible scores range from 0 to 100, with higher scores indicating better health status. We will calculate and present the physical component summary score and its associated domain scores (physical functioning, role-physical, bodily pain, general health).

##### 10-year cardiovascular risk

We will calculate the 10-year risk of myocardial infarction, stroke or cardiovascular death using three risk assessment tools. We will use one of the three risk assessment tools for each individual patient based on their disease profile: The SMART tool for patients with cardiovascular disease (35), the ADVANCE tool for patients with type 2 diabetes and no cardiovascular disease (36), and the SCORE2 tool for patients with no cardiovascular disease or type 2 diabetes (37). All data needed for the risk assessment tools are already being collected as routine practice. Results derived from the three different risk assessment tools will be pooled as they measure the exact same outcome (10-year absolute cardiovascular risk).

#### Additional outcomes

##### Body weight

We will measure the patients’ body weight and present it as mean percentual change in kg.

##### Waist circumference

We will measure waist circumference with a tape measurer and present it as mean change in cm.

##### Health-related quality of life – mental

We will administer the 36-Item Short Form Survey questionnaire for all patients (33). Possible scores range from 0 to 100, with higher scores indicating better health status. We will calculate and present the mental component summary score and its associated domain scores (vitality, social functioning, role-emotional, mental health).

##### Weight bias internalization

We will administer the Weight Bias Internalization Scale (WBIS) questionnaire for all patients (38). The WBIS includes 19-items to be rated on a 7-point Likert scale ranging from “strongly disagree” to “strongly agree”. We will convert the answers so that higher scores indicate greater weight bias internalization and present them as mean scores of all 19 items.

##### Cardiorespiratory fitness

We will measure maximal oxygen uptake (VO_2_max) with the Seismofit® System (VentriJect ApS, Hellerup, Denamark) (39). Cardiovascular fitness is considered a crucial risk factor for cardiovascular morbidity and death (40). The Seismofit® sensor is placed on the sternum, where it measures seismocardiography (the vibrations generated by the heart and transmitted through the chest). These measures are coupled with information on sex, age, weight, and height. Using an algorithm, the Seismofit® System then estimates VO_2_max.

##### Lower body strength

We will measure lower body strength with the 30-Seconds Sit-To-Stand Test (41). The 30-Seconds Sit-to-Stand test has been widely used in populations with obesity and is considered both safe and reproducible (42).

##### Walking capacity and endurance

We will measure walking capacity and endurance with the 6-Minute Walk Test. The 6-Minute Walk Test is both valid and reliable in populations with obesity (43,44).

## Study Procedures

### Recruitment

We will invite all eligible patients to participate in this study when they enter the weight loss program. At the first dietician consultation, one of the program’s two dieticians will briefly present the study, hand out written study information, and ask permission to contact them by phone in relations to the study. The primary investigator will then contact the patient by phone to provide further information about study participation. Should the patient wish to join the study, the first data collection visit will be scheduled either subsequently to the next dietician appointment a week later or at a preferred time within the program’s two first weeks. At this meeting, the patient will provide written informed consent and formally enter the study.

### Data Collection

We will collect data at three timepoints during the weight loss program: at the start of the program (baseline), at the early results evaluation visit (four months), and at end of the program (24 months). Data collection visits will take place right after the standard scheduled clinical visits at three timepoints during the program to ease participation burden and ensure timely data collection. The primary investigator will collect data at baseline and four months for all patients. At 24 months, data will be collected by a research assistant. Data collection visits at four and 24 months will be identical to the baseline visit, except that demographic information will be collected only once. All data will be collected using a tablet and stored directly on a secure electronic REDCap database. Similarly, consent forms will be uploaded to a secure electronic platform (“L-drevet”). See Table 1 for details of the data collection.

**TABLE 1:**
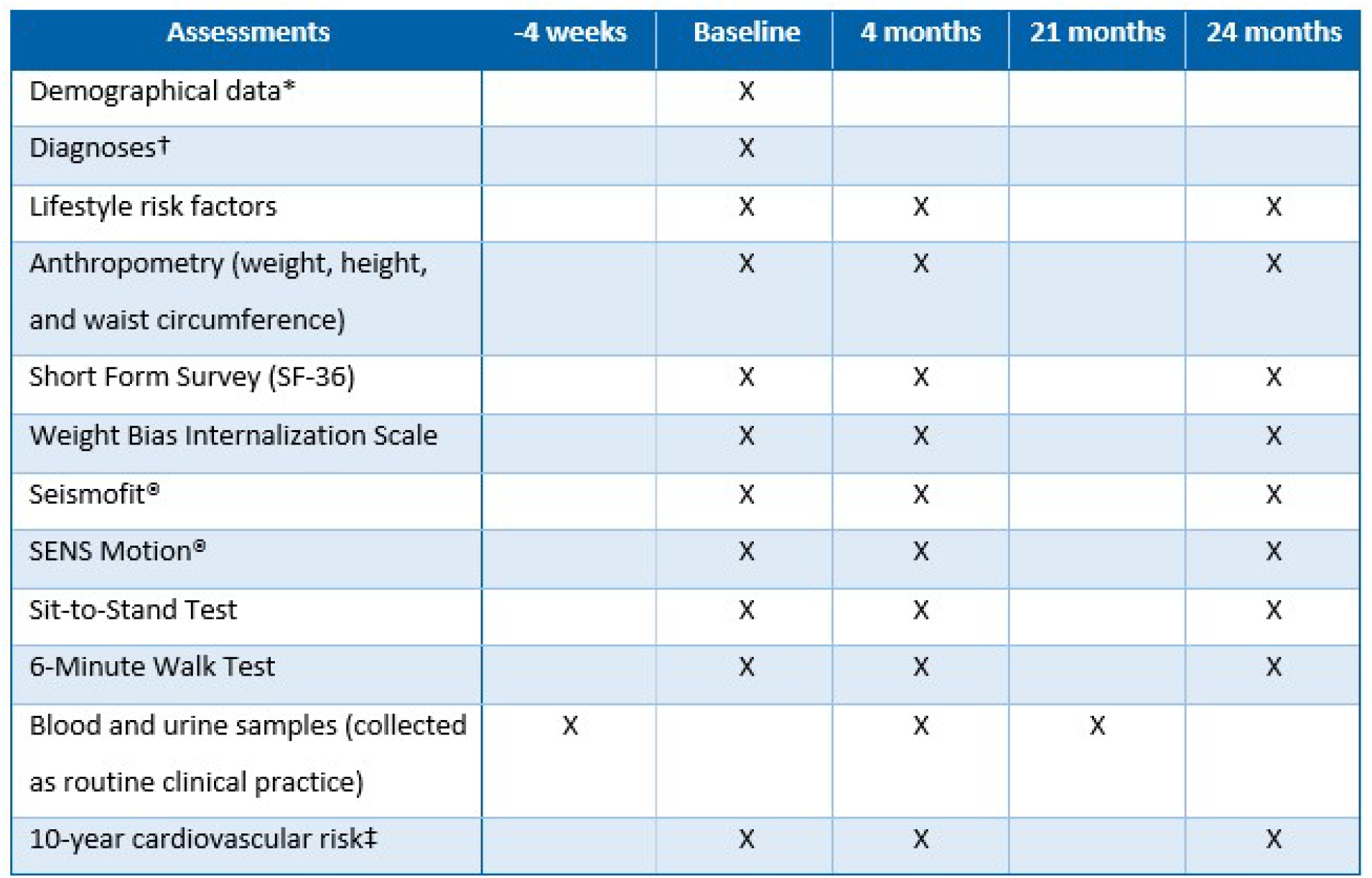
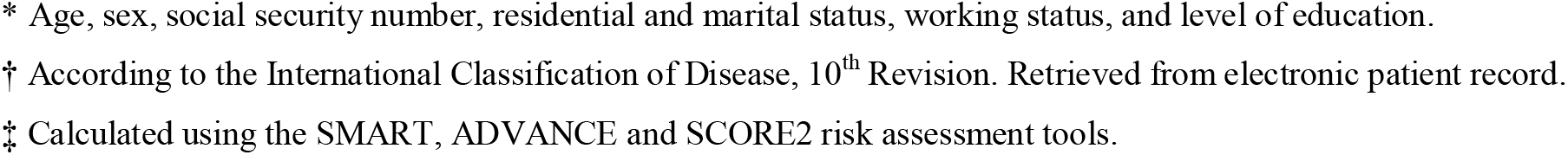
Overview of data collection.

| Assessments | -4 weeks | Baseline | 4 months | 21 months | 24 months |
| --- | --- | --- | --- | --- | --- |
| Demographical data* |  | X |  |  |  |
| Diagnoses† |  | X |  |  |  |
| Lifestyle risk factors |  | X | X |  | X |
| Anthropometry (weight, height, and waist circumference) |  | X | X |  | X |
| Short Form Survey (SF-36) |  | X | X |  | X |
| Weight Bias Internalization Scale |  | X | X |  | X |
| Seismofit® |  | X | X |  | X |
| SENS Motion® |  | X | X |  | X |
| Sit-to-Stand Test |  | X | X |  | X |
| 6-Minute Walk Test |  | X | X |  | X |
| Blood and urine samples (collected as routine clinical practice) | X |  | X | X |  |
| 10-year cardiovascular risk‡ |  | X | X |  | X |

The data collection visits will take place in a clinical investigation room. Firstly, we will collect demographical information, conduct anthropometrical measurements (weight, height, and waist circumference), and administer the questionnaires (SF-36, WBIS-19). Next, the patient will lie supine on a bed and rest while we use the Seismofit® system to assess VO_2_max. Then, we will attach the SENS Motion® sensor on the participant’s thigh, laterally 10 cm above the knee. We will ask them to wear it underneath normal clothing for seven full days before removing it and mailing it to us in a padded envelope. Lastly, patients will perform the functional tests in the investigation room and in a nearby hallway.

Immediately after the data collection visits, we will access the patient records to retrieve blood and urine test results that are specifically necessary for the risk assessment tools (serum creatinine, cholesterol, glycated hemoglobin, c-reactive protein, and albumin/creatinine ratio). These test results are already collected as part of clinical practice prior to the two physician consultations four weeks before program start and at four months. However, for the final data collection visit, blood and urine samples are collected already at 21 months. This means that data for the 10-year cardiovascular risk will be assessed at 21 months while the remaining data collection occurs at the actual end of the program at 24 months.

### Patient and Public Involvement

Patients and healthcare professionals were involved in several aspects of the preparation of this study. To guide us in selecting outcomes, we informally interviewed nine patients about how they hoped the program would benefit them and other patients, and how we might best measure that. Likewise, we interviewed two endocrinologists working with the program about which outcomes would hold the most value for clinical decision-making. This resulted in the selection of three key outcome measures with particular interest for patients and physicians, namely physical activity, SF-36 physical component, and 10-year cardiovascular risk. Additionally, we pilot tested the selected functional tests (Sit-to Stand Test and 6-Minute Walk Test) with 10 patients and received feedback to ensure that tests and data collection procedures were feasible and acceptable for this particular patient population.

### Sample Size Considerations

Most exploratory studies are not designed to test hypotheses and stipulate about causal inferences, which is why they do not commonly have a classical approach to sample size estimation (45). It is recommended, however, that the sample size is justified in some way. The sample size of the present study was determined pragmatically and mainly based on available time and resources. We deem it feasible to recruit patients for a six-month period. With an expected inclusion of 75 patients annually in the clinical weight loss program, we aim to recruit 35 patients for this study. Based on our current observations, we expect a dropout rate of approximately 20 % (a combination of patients excluded at the early results evaluation and random dropouts during the program), resulting in an estimated sample size of 28 patients by the end of the program. We plan to contact the potential dropouts to explore their reasons for dropping out.

### Statistical Analysis

We will use descriptive statistics for patient demographics and baseline measurements of all outcomes and present them as numbers with percentages, means with standard deviations, or medians with interquartile ranges, depending on the type and distribution of data. Similarly, we will use descriptive statistics and no hypothesis testing to assess changes in outcomes during the weight loss program. We will also explore whether changes occur during the initial period of the program (between baseline and four months) or the remainder of the program (between four months and 24 months). All outcomes will be reported and presented as mean changes with 95 % confidence intervals from baseline to 24 months, baseline to four months, and four months to 24 months. For outcomes, whose change scores cannot be assumed to have normal distribution, we will bootstrap 95 % confidence intervals.

### Study Timeline

The study will run from primo 2024 to medio 2027. Data collection began February 2024, as stated in the NCT-registration. See Table 2 for a detailed Gantt chart of the study.

**TABLE 2:**
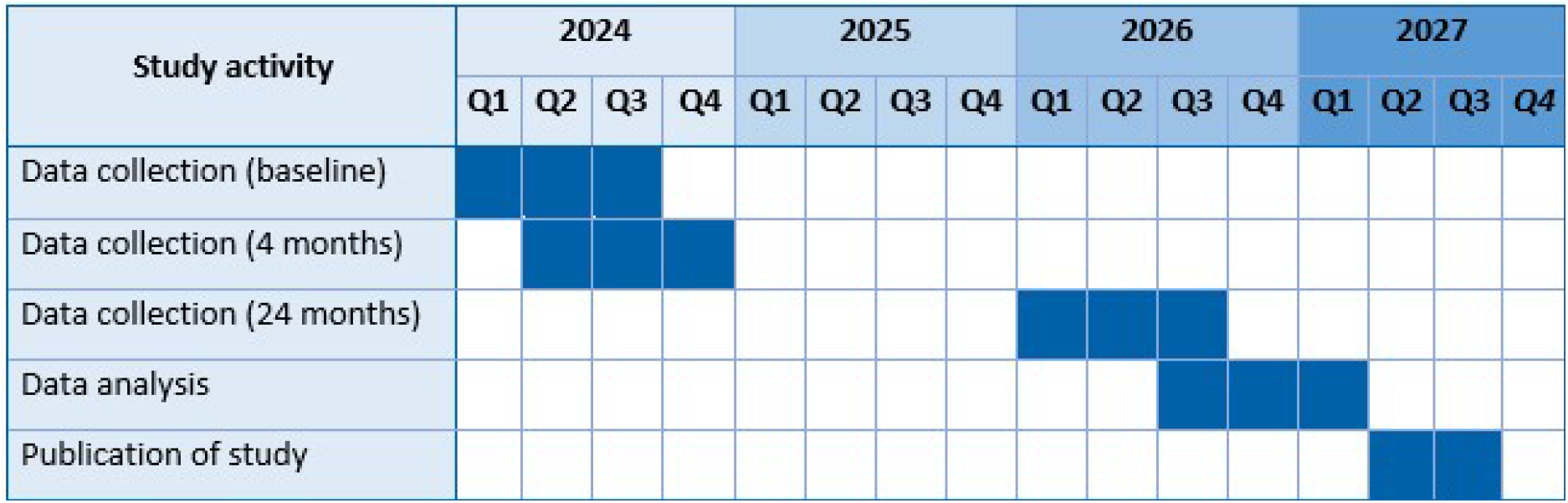
Gantt chart of study timeline.

### Ethics

The project has been approved to collect and save data by the Capital Region of Denmark’s legal administrative service “Privacy” (journal number: p-2023-15245). All study participants are presented both verbal and written study information, clearly stating that participation in this study is not a requisite to be in the weight loss program. After having the study explained, they are given ample time to consider participation before signing the informed consent. Data collection procedures have been pilot tested to ensure minimal participation burden, especially in regard to the physical tests. Furthermore, the method for assessing physical activity (accelerometry sensors) is considered both safe and without risks. At worst, skin irritation might occur due to the adhesive tape in which case the sensor can easily be removed entirely. Thus, we do not consider this study to be associated with any health research ethical issues. Nonetheless, we presented the project plan to the Capital Region’s Research Ethics Committee who determined, that it does not require formal approval (F-23061071). Finally, the study applies with the Declaration of Helsinki.

### Dissemination Plan

We plan to disseminate the findings of the project both written and verbally. Dissemination activities include oral and poster presentations at academic symposia and conferences, social media, and presentations to clinical end-users. Furthermore, we will publish the findings of this prospective cohort study as well as other planned SemPAct project studies in international peer-reviewed journals (working titles outlined below). We expect to publish at least three articles. Authorship is granted in accordance with the Vancouver Guidelines.

### Preliminary Publication Plan

#### Tentative title of prospective cohort study

*Exploring physical capacity, physical activity, and quality of life in patients with severe obesity and multimorbidity during an intensive Semaglutide weight loss program: A real-life prospective cohort study*.

Authors: Rasmus H. Brødsgaard, Carsten Dirksen, Mette M. Pedersen, Jeanette W. Kirk, Olivia M. Duun, Minna Titika Finken, Kirstine N. Bojsen-Møller^*^, Thomas Bandholm^*^ (^*^sharing last authorship).

#### Tentative title of interview study

*Understanding Physical Activity for Individuals with Obesity and Multimorbidity in an Intensive Weight Loss Program: An Interview Study with Patients and Healthcare Professionals*.

Authors: Rasmus H. Brødsgaard, Carsten Dirksen, Kirstine N. Bojsen-Møller, Thomas Bandholm, Jeanette W. Kirk.

#### Tentative title of co-design study

*Co-designing exercise for multi-disciplinary Semaglutide-centred weight loss treatment*.

Authors: Rasmus H. Brødsgaard, Carsten Dirksen, Kirstine N. Bojsen-Møller, Thomas Bandholm, Jeanette W. Kirk.

## Data Availability

All data produced in the present study will be available upon reasonable request to the authors.

